# Plasma Proteogenomic Uncovers Novel Molecular Mechanisms of Soluble Endoglin and Identifies AZI2 as a new Player in Arteriovenous Malformations in Hereditary Hemorrhagic Telangiectasia

**DOI:** 10.64898/2026.11.18.26366478

**Authors:** Gaëlle Munsch, Carole Proust, Catherine E. Richter, Marine Germain, Maud Tusseau, Alexandre Guilhem, Shirine Mohamed, Charlotte Richardot, Floriane Samaria, Florian Thibord, Ilana Caro, Claire Bardel, Eike Struck, INVENT consortium, Robert Olaso, Jean-François Deleuze, Stéphanie Debette, Franck Peiretti, Lynn M Butler, Béatrice Jaspard-Vinassa, Jordan A. Shavit, Elisa Rossi, Sophie Dupuis-Girod, David-Alexandre Trégouët, Omar Soukarieh

## Abstract

Endoglin is a transmembrane glycoprotein that plays a key role in vascular development, angiogenesis and endothelial homeostasis. Its soluble form (Sol-ENG), generated through proteolytic shedding of the membrane-bound protein, has been implicated in the pathophysiology of numerous cardiovascular diseases, ranging from common conditions such as preeclampsia and thrombosis to rare diseases such as hereditary hemorrhagic telangiectasia (HHT).

Leveraging large-scale genomics, transcriptomics, proteomics and metabolomics data from public and original resources, we identify a molecular signature of 24 markers associated with plasma levels of Sol-ENG. This signature was enriched in genes coding for GPI-enchored proteins and included metabolites participating to glycosylation and redox homeostasis. Furthermore, we observed that ABO blood groups and genetically predicted plasma levels of 5-Azacytidine Induced 2 (AZI2) associate with the risk of arteriovenous malformations in HHT patients.

This study provides new insights into the physiological mechanisms underlying Endoglin regulation and opens clinical perspectives for the management of HHT patients.

## Introduction

Endoglin is a membrane glycoprotein that plays a major role in endothelial cell biology and is highly involved in angiogenesis (Rossi et al. 2019; López-Novoa et Bernabeu 2010). Endoglin has mainly been known as co-receptor in response to bone morphogenetic proteins belonging to the transforming growth factor β (TGF-β) superfamily (Nolan-Stevaux et al. 2012; Lebrin et al. 2004; Saito et al. 2017). A membrane bound and a soluble form (Sol-ENG) of Endoglin have been described, the latter mainly resulting from a protease cleavage of the former (Bellón et al. 1993; Hawinkels et al. 2010).

For many years, studies on Endoglin have concentrated on membrane-bound Endoglin and its role in Hereditary Hemorrhagic Telangiectasia (HHT), a multiorgan rare vascular disease, arising in ~60% cases from Endoglin deficiency due to loss-of-function (LoF) mutations in the *ENG* gene, encoding for Endoglin. HHT is characterized with very heterogeneous clinical manifestations between patients, thus requiring a multidisciplinary approach for the clinical diagnosis and treatment (Ola et al. 2023; Kritharis et al. 2018). The clinical diagnosis of HHT is based on Curaçao criteria corresponding to epistaxis, multiple telangiectasia at characteristic sites (lips, oral cavity, fingers and nose), visceral vascular lesions including gastrointestinal telangiectasia and/or arteriovenous malformations (AVMs), and family history (Faughnan et al. 2020). Frequent and repeated nose bleeding and/or digestive telangiectasia can lead to anemia and visceral AVMs lead to vascular shunts, strokes and cerebral abscess secondary to pulmonary AVMs and high cardiac output secondary to liver AVMs (Faughnan et al. 2020). Sol-ENG has also been proposed as a possible biomarker for HHT as reduced levels of the Sol-ENG have been observed in plasma of HHT patients carrying *ENG* LoF variants (Botella et al. 2015; Ojeda-Fernandez et al. 2010). Increased Sol-ENG has also been associated with a reduced number of AVMs in a HHT mouse model, suggesting that sol-ENG could be involved in HHT and its complications (Gallardo-Vara et al. 2018).

More recent works have highlighted new functions for Endoglin (Rossi et Bernabeu 2023), in particular in leukocyte adhesion and transmigration under inflammatory conditions (Rossi et al. 2013) but also in platelets biology (Rossi et al. 2023) and cancer-associated fibroblasts (Paauwe et al. 2018). There is then accumulating evidence that Endoglin and its soluble form are involved in many human diseases (Bernabeu et al. 2023) such as thrombosis (Rossi et al. 2023), coronary atherosclerosis (H. Chen et al. 2021), preeclampsia (Margioula-Siarkou et al. 2022), hypertension (Blázquez-Medela et al. 2010), autoimmune disease (Grignaschi et al. 2022), some cancers (Hakuno et al. 2023), and beyond.

In this work, we hypothesized that elucidating the molecular determinants of Sol-ENG could deepen our understanding of the pathophysiology underlying complex traits associated with Endoglin and may also improve the clinical management of vascular complications in HHT patients. To address this hypothesis, we leveraged results from large-scale plasma proteogenomics studies on Sol-ENG plasma levels and transcriptomics data from monocytes and endothelial cells measuring Endoglin mRNA levels. This approach allowed us to identify key molecular determinants and assess their clinical impact on the risk of AVMs in a cohort of HHT patients.

## Results

### Common polymorphisms associate with Endoglin plasma levels

By meta-analyzing results from two genome wide association studies (GWAS) on Sol-ENG plasma levels measured using an aptamer-based SomaScan technology in 46,267 healthy individuals, we identified eight genomic regions harboring single nucleotide polymorphisms (SNPs) significantly (p<5×10^−8^) associated with Sol-ENG plasma levels (**Figure 1, Supplemental Table 1**). Clumping analysis with pairwise linkage disequilibrium (LD) r^2^<0.01 further revealed 24 independent SNPs associated with Sol-ENG levels over these eight regions (**Table 1**). The association of seventeen of these SNPs with Sol-ENG replicated (p<4×10^−3^, unilateral corrected p-value after Bonferroni correction) in the UK Biobank population where Sol-ENG was measured using an Olink technology (**Table 1**). Of note, using the summary statistics of the GWAS on Sol-ENG based on the SomaScan and Olink approaches, we estimated that the genetic correlation between these two measurements of Sol-ENG was 0.88 ± 0.17 (p=1.96×10^−7^), providing support using UK Biobank as a validate tool to replicate the observed associations.

**Table 1.** Independent single nucleotide polymorphisms significantly associated at p<5×10^−8^ with Sol-ENG in the metaGWAS analysis and their replication in UK Biobank.

| Chromosome region | Variant-EA/NEA | Position (Hg38) | (Nearest) Gene | metaGWAS (N=46,267) | UK Biobank (N=34,557) |
|---|---|---|---|---|---|
| 6q23.3 | rs1547247-A/G* | 135069698 | HBS1L;MYB | 0.04 ± 0.01 ; P=1.98x10<sup>-8</sup> | 0.02 ± 0.01 ; P=2.12x10<sup>-3</sup> |
| 9q33.3 | rs11998752-C/G | 124452866 | PSMB7;NR5A1 | 0.09 ± 0.02 ; P=1.08x10<sup>-8</sup> | -0.01 ± 0.02 ; P=0.779 |
|  | rs10819187-A/G | 126607692 | MVB12B;LMX1B | -0.05 ± 0.01 ; P=4.59x10<sup>-8</sup> | 0.02 ± 0.01 ; P=0.138 |
| 9q34.11 | rs72752823-A/G | 127734901 | TOR2A | -0.06 ± 0.01 ; P=3.33x10<sup>-10</sup> | -0.02 ± 0.01 ; P=0.119 |
|  | rs1054775-A/G* | 127793005 | CDK9;FPGS | -0.05 ± 0.01 ; P=2.15x10<sup>-13</sup> | -0.05 ± 0.01 ; P=2.37x10<sup>-10</sup> |
|  | rs10987756-C/G* | 127849244 | ENG | 0.14 ± 0.01 ; P=2.35x10<sup>-43</sup> | 0.28 ± 0.01 ; P=9.08x10<sup>-105</sup> |
| 9q34.13 | rs141524184-T/C | 132877425 | AK8 | -0.19 ± 0.03 ; P=2.80x10<sup>-8</sup> | -0.02 ± 0.04 ; P=0.561 |
|  | rs117405640-A/G* | 133015880 | GFI1B;SNORD141A | -0.42 ± 0.03 ; P=1.93x10<sup>-57</sup> | -0.49 ± 0.03 ; P=9.20x10<sup>-52</sup> |
|  | rs34813452-A/G* | 133160728 | GBGT1 | -0.08 ± 0.01 ; P=2.68x10<sup>-10</sup> | -0.09 ± 0.01 ; P=4.75x10<sup>-10</sup> |
|  | rs10793956-A/G* | 133204155 | GBGT1;OBP2B | 0.18 ± 0.02 ; P=7.53x10<sup>-16</sup> | 0.21 ± 0.03 ; P=5.93x10<sup>-17</sup> |
|  | rs75212266-A/G* | 133207463 | OBP2B | -0.26 ± 0.03 ; P=3.16x10<sup>-20</sup> | -0.19 ± 0.03 ; P=6.63x10<sup>-11</sup> |
| 9q34.2 | rs558240-A/G* | 133281560 | ABO;SURF6 | -0.24 ± 0.01 ; P=4.25x10<sup>-262</sup> | -0.27 ± 0.01 ; P=1.04x10<sup>-263</sup> |
|  | rs11523134-A/T* | 133513398 | SLC2A6;MYMK | 0.10 ± 0.01 ; P=2.93x10<sup>-33</sup> | 0.12 ± 0.01 ; P=3.79x10<sup>-31</sup> |
|  | rs17150674-T/C* | 133545810 | ADAMTSL2 | 0.10 ± 0.01 ; P=1.17x10<sup>-13</sup> | 0.14 ± 0.02 ; P=6.88x10<sup>-18</sup> |
|  | rs118117764-A/G | 133785359 | VAV2 | -0.09 ± 0.01 ; P=4.07x10<sup>-12</sup> | -0.01 ± 0.02 ; P=0.757 |
| 11q24.2 | rs8177398-T/C* | 126287999 | TIRAP | 0.18 ± 0.02 ; P=8.82x10<sup>-14</sup> | 0.12 ± 0.02 ; P=5.12x10<sup>-7</sup> |
|  | rs10893497-T/C* | 126330641 | DCPS | -0.06 ± 0.01 ; P=1.49x10<sup>-12</sup> | -0.04 ± 0.01 ; P=8.19x10<sup>-6</sup> |
|  | rs7113465-T/C* | 126424193 | KIRREL3 | 0.04 ± 0.01 ; P=2.14x10<sup>-8</sup> | 0.04 ± 0.01 ; P=6.92x10<sup>-8</sup> |
| 17p13.1 | rs67143157-A/G* | 7171399 | ASGR2;ASGR1 | -0.04 ± 0.01 ; P=1.83x10<sup>-8</sup> | -0.06 ± 0.01 ; P=4.78x10<sup>-13</sup> |
| 19q13.2 | rs284660-T/G* | 41426179 | B3GNT8 | 0.04 ± 0.01 ; P=8.46x10<sup>-10</sup> | 0.03 ± 0.01 ; P=1.17x10<sup>-5</sup> |
| 19q13.31 | rs117498036-A/G | 43429736 | TEX101;LYPD3 | -0.10 ± 0.02 ; P=9.05x10<sup>-9</sup> | -0.05 ± 0.02 ; P=7.63x10<sup>-3</sup> |
|  | rs1055099-T/G | 43475437 | PHLDB3 | 0.04 ± 0.01 ; P=1.57x10<sup>-8</sup> | 0.01 ± 0.01 ; P=0.268 |
|  | rs4760-A/G* | 43648948 | PLAUR | -0.26 ± 0.01 ; P=1.14x10<sup>-158</sup> | -0.24 ± 0.01 ; P=3.09x10<sup>-131</sup> |
| 20q11.22 | rs73106997-T/C* | 34787335 | NCOA6 | -0.06 ± 0.01 ; P=1.12x10<sup>-12</sup> | -0.03 ± 0.01 ; P=1.23x10<sup>-3</sup> |

EA: Effect Allele; NEA: Non Effect Allele; \*Associations that replicate at p<4x10<sup>-3</sup> in UK Biobank

**Figure 1.**
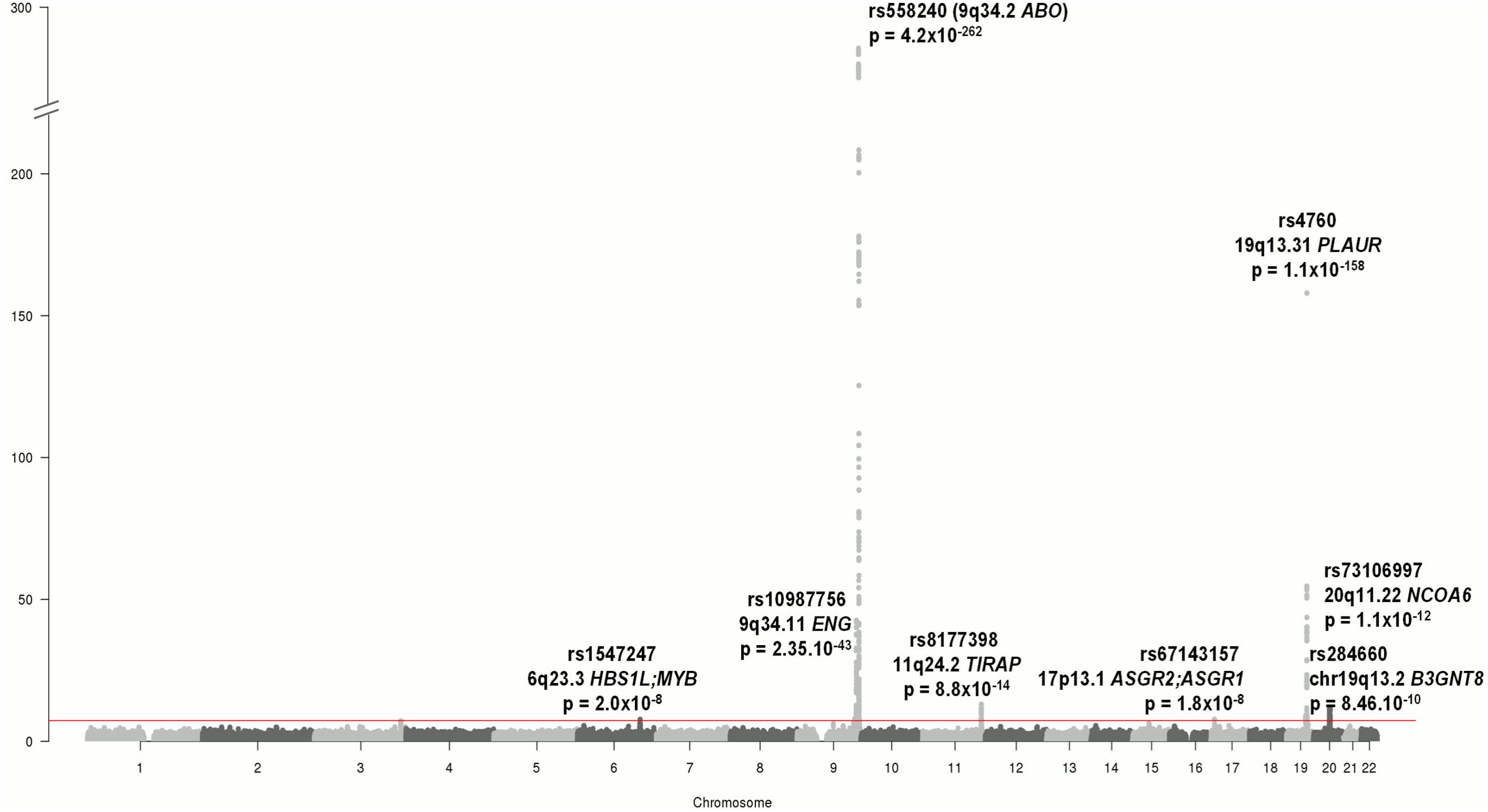
Manhattan plot summarizing the meta-analysis of two GWAS datasets on plasma Endoglin levels in 46,091 individuals. The identity, location and p-value of the eight loci significantly (p<5×10^−8^) associated with Endoglin levels are indicated. The exact genomic position (in GRCh38) of the lead SNPs are: chr6:135069698 (*HBS1L/MYB* rs1547247), chr9:127849244 (*ENG* rs10987756), chr9:133281560 (*ABO* rs558240), chr11:126287999 (*TIRAP* rs8177398), chr17:7171399 (*ASGR2;ASGR1* rs67143157), chr19:41426179 (*B3GNT8* rs284660), chr19:43648948 (*PLAUR* rs4760) and chr20:34787335 (*NCOA6* rs73106997).

As pairwise r^2^ is a suboptimal metric to fully characterize the extend of LD at a given locus, we conducted further LD and haplotype analyses at the three genomic regions presenting several replicated Sol-ENG associated SNPs after clumping: 9q34.11 (*CDK9-ENG*), 9q34.2 (*GFI1B-GBGT1-OBP2B-ABO-SURF6-SURF6-ADAMTSL2*) and 11q24.2 (*TIRAP-DCPS-KIRREL3*).

While at 9q34.11 the rs105477 (*CDK9*) and rs10987756 (*ENG*) were not in LD (D’=0.03, r^2^=0.00) and therefore considered as two independent signals, the *TIRAP* rs8177398 was in strong negative LD (D’=−1) with the *DPCS* rs10893497 and *KIRREL3* rs7113465 at the 11q24.2 locus. Haplotype analysis of the latter three SNPs in 966 participants of the 3C study strongly suggests that the rare haplotype carrying the *TIRAP* rs8177398-T allele is mainly responsible for the association signal observed with Sol-ENG at the 11q24.2 locus (**Supplemental Table 2**).

At 9q34.2 where the most significant association was observed in the metaGWAS analysis, the lead SNP was rs558240. This variant is in strong LD with the *ABO* variants that characterize the ABO blood groups: rs2519093 (A1), rs1053878 (A2), rs8176743 (B), rs8176719 (O1) and rs41302905 (O2) (Goumidi et al. 2021). In particular, the rs558240-A allele is either present on the A1 haplotype or subdivides the O1 groups into two subhaplotypes. It is never associated with the B, O2 nor A2 blood groups (**Supplemental Figure 1**). To further clarify the association observed at the *ABO* locus, we investigated the association of ABO blood groups with Sol-ENG plasma levels in the 3C study (Godin et al. 2008). The pattern of associations is shown in **Figure 2**. To summarize, in this healthy population, assuming additive allele effects, the ABO B group was associated with increased Sol-ENG plasma levels (β=+0.49 ± 0.076, p=1.87×10^−10^) compared to the O1 blood group whereas the A1 group was associated with decreased levels (β=−0.25 ± 0.048, p=2.94×10^−7^). After adjusting for the B and A1 effects, the association p-value of rs558240 with Endoglin decreased from p=1.02×10^−6^ to p=0.439. Altogether, ABO blood groups explained ~6% of the inter-individual variability in Sol-ENG plasma levels in the 3C study. Further LD and regression analyses adjusted for ABO blood groups conducted in the 3C study strongly suggest that the association of other 9q34.2 SNPs with Sol-ENG is likely due to LD with ABO blood groups (**Supplemental Table 3 & Supplemental Table 4**).

**Figure 2.**
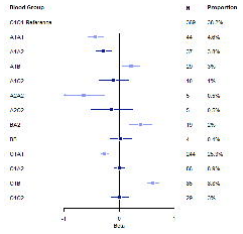
Forest plot summarizing the association of ABO diplotypes with Endoglin plasma levels. Diplotype effects were assessed with the O1O1 diplotype as the reference and were estimated based on standardized Endoglin values measured in 966 participants from the 3C study, where Endoglin levels were determined using Olink technology. N, number of carriers.

In summary, these investigations of the main meta-GWAS findings led to the identification of nine independent loci associated with Sol-ENG. In addition to ABO blood groups (with rs558240 as lead SNP), the lead SNPs at these loci, ordered by effect size, are: *PLAUR* rs4760 (19q13.31), *TIRAP* rs8177398 (11q24.2), *ENG* rs10987756 (9q34.11), *NCOA6* rs73106997 (20q11.22), *CDK9* rs1054775 (9q34.11), *ASGR1* rs67143157 (17p13.1), *B3GNT8* rs284660 (19q13.2) and *HBS1L/MYB* rs1547247 (6q23.3).

Altogether, the ABO blood groups and the eight other lead Sol-ENG associated variants explained ~9.5% of the interindividual variability of Sol-ENG plasma levels in the 3C Study.

### Transcriptome-Wide Association Study (TWAS) identifies novel loci participating to sol-ENG regulation

–TWAS identified 434 inferred gene expressions across various tissues, over 61 unique genes, that were significantly (P<8.3×10^−6^) associated with Sol-ENG plasma levels (**Supplemental Table 5, Supplemental Figure 2**). From these associations, 101 mapping to 26 unique genes demonstrated a high colocalization PP4 probability (PP4>0.75). Finally, a conditional analysis provided evidence for 24 unique genes as involved in Sol-ENG regulation. Among these 24 genes, two genomic regions were novel (**Figure 3**) in the sense that they were not detected in the metaGWAS as none of their SNPs has individually reached the p<5×10^−8^ statistical threshold:

**Figure 3.**
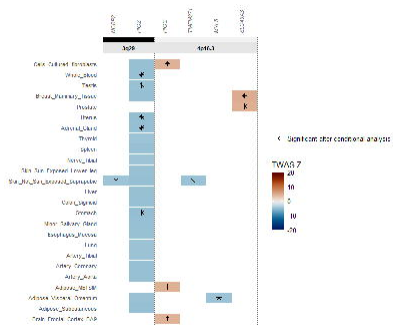
Heatmap summarizing the conditional TWAS and colocalization analyses at the two newly identified genomic region.

On chr3q29, increased expression of *NCBP2* and *PIGZ* in several tissues were associated with decreasing Sol-ENG levels. In the GWAS meta-analysis of Sol-ENG, the lead variant in this region was the synonymous *PIGZ* rs12636891 (p.Leu297Leu) where the G allele was associated with decreased Sol-ENG plasma levels (β=−0.029 ± 0.007, p=9.31×10^−6^). This variant does not exhibit any LD (r^2^>0.80) with any other SNP. Of note, the association of *PIGZ* rs12636891-G allele with Sol-ENG levels showed the same direction in UK Biobank (β=−0.012 ± 0.007), but failed to achieve statistical significance for replication (p=0.083).

On chr4p16.3, expression of four genes showed significant association with Sol-ENG according to the studied tissues: *PIGG, TMEM271, MYL5* and *SLC49A3*. The lead variant was the downstream *PIGG* rs77060885 with the T allele associated with increased Sol-ENG plasma levels (β=0.078 ± 0.019, p=4.40×10^−5^). Of note, the association of rs77060885-T allele with Sol-ENG plasma levels in the UK Biobank was also borderline (β=0.036 ± 0.020, p=0.081).

The TWAS analysis also enabled us to refine some of the association signals that were detected in the metaGWAS analysis (**Supplemental Figure 2**). In particular, while *B3GNT8* was identified in the metaGWAS as the lead locus on chr19, the TWAS driven framework suggested that the *EXOSC5, BCKDHA, DMAC2* and *ERICH4* could be as relevant as *B3GNT8* to explain the association signal at 19q13.2 since these four gene expressions were selected after conditional analyses on other gene expressions. In particular, for lung tissue, *BCKDHA* gene expression was kept after conditional analysis on *B3GNT8* gene expression. Similarly, on 20q11.22, the conditional analysis identified *PIGU* or *GSS* as the most plausible gene instead of the *NCOA6* locus identified in the GWAS. Of note, the lead variant at *PIGU* in the metaGWAS for Sol-ENG was rs6088537 (p=1.73×10^−12^). It is in strong LD (r^2^=0.97) with the best eQTL (rs6088525) for *PIGU* expression and with the lead *NCOA6* rs73106997 variant (r^2^=0.37, D’=+0.98)

Overall, the TWAS analysis identified new biological candidates, in particular by highlighting three genes, *PIGU, PIGZ* and *PIGG*, belonging to the family of genes involved in the biosynthesis of glycosylphosphatidylinositol (GPI) anchors. As shown in **Supplemental Table 5**, most of these TWAS findings replicated in UK Biobank.

### Plasma protein and metabolite based Mendelian Randomization (MR) approaches

MR analysis leads to the identification of nine proteins (AZI2, BGAT, CTRC, FIX, LGALS4, NOTCH1, SELE, SERPINI2 and TNFRSF10C) with genetically determined plasma levels significantly associated with Sol-ENG levels (**Supplemental Table 6**). Two of them, 5-Azacytidine Induced 2 (AZI2) and coagulation factor IX (FIX), exhibited no statistical evidence for pleiotropy nor reverse causality, with increased plasma levels of both proteins associated with decreased Sol-ENG. Genetic instruments used in UK Biobank to infer genetically determined plasma levels of these two proteins are given in **Supplemental Table 7**. Of note, none of these genetic instruments are cis-acting pQTL variants.

Besides, plasma levels of four metabolites were observed to be associated with Sol-ENG levels without evidence for reverse causality (**Supplemental Table 8**). Genetically-determined plasma levels of Asparte to mannose ratio and Glucose-to-mannose ratio were associated with decreased Sol-ENG while genetically determined Gamma-glutamylglycine and Glycine levels were positively associated with plasma Sol-ENG. The analysis of their genetic instruments (**Supplemental Table 9)** showed that Gamma-glutamylglycine and Glycine shared genetic regulators (*ACADM, ALDH1L1-AS2, CSP1* and *GLDC*) while *PPMG1* is a common regulator of both the Aspartate to mannose and Glucose-to-mannose ratios. All of these metabolites are indicators of the cell’s overall metabolic state, in particular energy status (glucose, mannose), glycosylation (glucose, mannose) and redox homeostasis and detoxification (glycine, gamma-glutamylglycine).

### Impact of identified molecular phenotypes on ENG expression in monocytes and endothelial cells

We then examined whether the expression of TWAS-identified genes and genes encoding MR-identified proteins correlate with *ENG* expression in the two primary cell types in which Endoglin plays key roles: endothelial cells and monocytes (Fernandez-L et al. 2006). Results are shown in **Supplemental Table 10**. Notably, *AZI2, NCOA6* and *PIGU* expressions consistently showed a negative correlation with *ENG* expression in both cell types. We also leveraged the genome wide genotype data available in 758 participants of the Cardiogenics Transcriptomic Study (CTS) (Garnier et al. 2013) to investigate whether the lead SNPs associated with Sol-ENG plasma levels were associated with *ENG* mRNA levels in monocytes. The *ENG* rs10987756-C allele was significantly (β=0.88 ± 0.08, p=4.45×10^−24^) associated with reduced *ENG* transcript levels (**Supplemental Table 11**) and accounted for ~12% of the variability in monocyte *ENG* expression. None of the other eight pQTLs showed statistical evidence for association with monocyte *ENG* expression after controlling for the number of tested SNP (all p>0.006 = 0.05/9).

### Protein quantitative trait locus analysis reveals potential shared biological pathways involving Sol-ENG

We leveraged GWAS results from the Fenland, deCODE and UK Biobank proteogenomic studies to investigate whether the nine identified Sol-ENG associated SNPs also commonly influence plasma levels of other proteins beyond Sol-ENG. Detailed results of these investigations are reported in **Supplemental Table 12** and **Supplemental Table 13**. While plasma levels of no other proteins, beyond Sol-ENG, were influenced by all nine studied variants, seven of them affected plasma levels of Interleukin-1 receptor type 1. Ten proteins were observed to shared four Sol-ENG associated variants, including two cytokine receptor subunits involved in hematopoiesis and immune cell development (Interleukin-3 receptor subunit alpha, Interleukin-7 receptor subunit alpha) and four proteins participating to endothelial cell biology (E-Selectin, Hematopoietic progenitor cell antigen CD34, Platelet endothelial aggregation receptor 1, Thrombospondin type-1 domain-containing protein 1).

Several proteins participating to the placenta growth factor activity were also identified as sharing at least two pQTLs, including Angiopoietin-1 receptor, Ephrin type-A receptor 4, Ephrin type-B receptor 4, Fibroblast growth factor receptor 1and Tyrosine-protein kinase receptor TYRO3. These observations are consistent with studies reporting elevated Sol-ENG levels in preeclampisa, a condition associated with impaired placental functions. Of note, these placenta-related proteins share the *ABO* and *ASGR1* variants as pQTLs, two variants exerting pleiotropic effects on about 300 proteins. In contrast, the *ENG* rs10987756 is a pQTL for only Endoglin (**Supplemental Table 13**).

Notably, in line with our TWAS findings, we observed that another GPI-anchor protein (CD109) shared three pQTLs with Sol-ENG, namely *ABO* rs558240, *ASGR1* rs67143157 and *PLAUR* rs4760 (**Supplemental Table 13**).

### Clinical implication of genomic findings

To further assess the clinical relevance of our genomic findings, we first conducted MR analyses to explore potential causal associations between increased sol-ENG plasma levels and various human diseases where sol-ENG has been proposed as a biomarker. These diseases correspond to preeclampsia (Tyrmi et al. 2023), systemic sclerosis (Jiang et al. 2021), myocardial Infarction (Hartiala et al. 2021), coronary artery disease (Aragam et al. 2022), stroke (Mishra et al. 2022) and venous thromboembolism (Thibord et al. 2022). After excluding outlier effects and accounting for the pleiotropic effects of ABO, no statistical evidence was found for a causal association between Sol-ENG and the studied diseases (see Methods – **Supplemental Table 14**).

We then examined the association of the nine lead Sol-ENG associated SNPs, the three PIG family variants identified in TWAS, and ABO blood group tagging SNPs with cerebral and pulmonary arteriovenous malformations (CAVM and PAVM, respectively) in a cohort of 676 HHT patients (**Supplemental Table 15**). Allele frequencies of the studied polymorphisms in HHT patients with or without AVMs are shown in **Supplemental Table 16**. Only ABO blood group-tagging SNPs were significantly (p<0.05) associated with AVMs. Compared to the O1 blood group, the A1 and O2 blood groups were associated with an increased risk of AVMs, Odds Ratio (OR) =1.52 (p=0.003) and OR=4.04 (p=0.0004), respectively (**Table 2**). A trend toward an increased risk of AVMs was also observed for B blood group (OR=1.40, p=0.15). No significant association was observed with A2 (p=0.86). These associations appeared stronger in patients carrying *ENG* (likely)-pathogenic variants compared to those with *ACVRL1* variants (**Table 3**) but were homogeneous in patients with CAVM or PAVM (**Supplemental Table 17**).

**Table 2.** Association of ABO blood groups-tagging SNPs with AVM in HHT patients. N, number of individuals.

|  | **All (N = 676)** | ***ACVRL1* variant carriers (N = 404)** | ***ENG* variant carriers (N = 234)** |
|---|---|---|---|
| **A1** | OR = 1.52 [1.15 - 2.01] ; p = 0.0034 | OR = 1.30 [0.85 - 2.00] ; p = 0.2322 | OR = 1.93 [1.15 - 3.23] ; p = 0.0130 |
| **A2** | OR = 1.04 [0.64 - 1.72] ; p = 0.8658 | OR = 0.96 [0.43 - 2.13] ; p = 0.9223 | OR = 0.90 [0.39 - 2.10] ; p = 0.8172 |
| **B** | OR = 1.40 [0.88 - 2.21] ; p = 0.1533 | OR = 1.31 [0.62 - 2.77] ; p = 0.4736 | OR = 1.44 [0.68 - 3.01] ; p = 0.3370 |
| **O2** | OR = 4.04 [1.88 - 8.70] ; p = 0.0004 | OR = 2.69 [0.94 - 7.67] ; p = 0.0641 | OR = 8.25 [1.06 - 64.5] ; p = 0.0443 |

Odds Ratio (OR) with their confidence interval in brackets were obtained after adjusting for age, sex and mutated gene (*ACVRL1* or *ENG*) when appropriate, under the assumption of additive genetic effects. Subgroup analyses by mutated gene were limited to HHT patients carrying likely pathogenic or pathogenic variants in ACVRL1 or ENG.

**Table 3.**
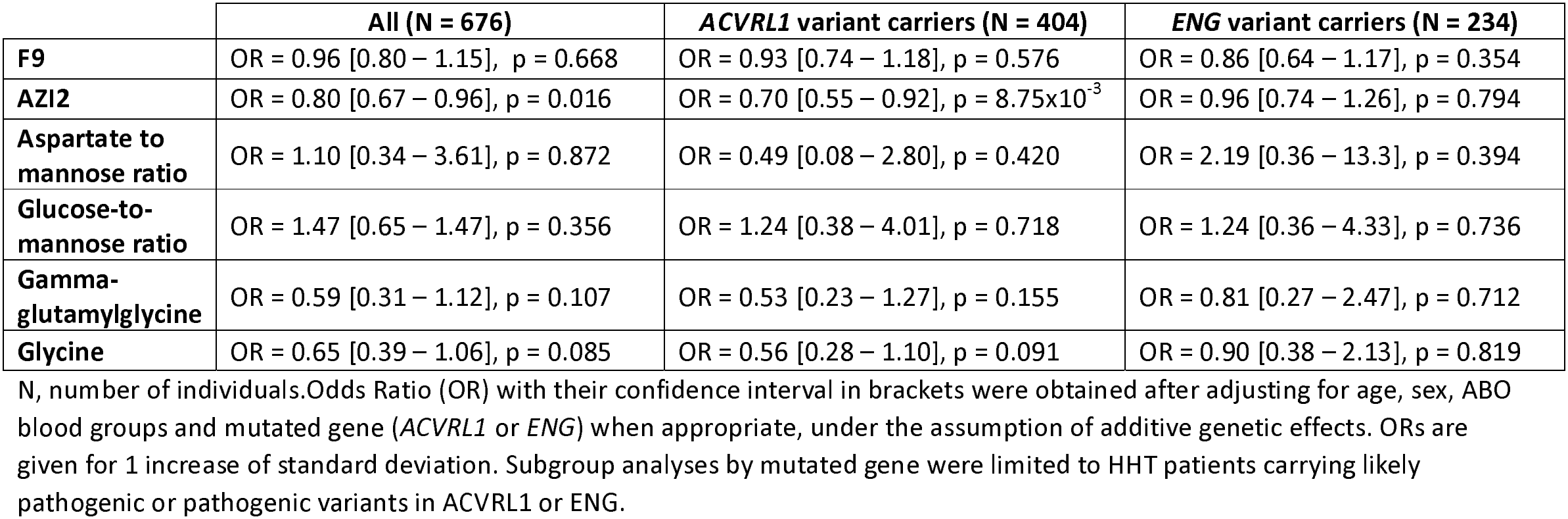
Association of MR-based genetically predicted plasma levels of candidate proteins/metabolites with AVM in HHT patients.

|  | **All (N = 676)** | ***ACVRL1* variant carriers (N = 404)** | ***ENG* variant carriers (N = 234)** |
|---|---|---|---|
| **F9** | OR = 0.96 [0.80 – 1.15], p = 0.668 | OR = 0.93 [0.74 – 1.18], p = 0.576 | OR = 0.86 [0.64 – 1.17], p = 0.354 |
| **AZI2** | OR = 0.80 [0.67 – 0.96], p = 0.016 | OR = 0.70 [0.55 – 0.92], p = 8.75x10<sup>-3</sup> | OR = 0.96 [0.74 – 1.26], p = 0.794 |
| **Aspartate to mannose ratio** | OR = 1.10 [0.34 – 3.61], p = 0.872 | OR = 0.49 [0.08 – 2.80], p = 0.420 | OR = 2.19 [0.36 – 13.3], p = 0.394 |
| **Glucose-to-mannose ratio** | OR = 1.47 [0.65 – 1.47], p = 0.356 | OR = 1.24 [0.38 – 4.01], p = 0.718 | OR = 1.24 [0.36 – 4.33], p = 0.736 |
| **Gamma-glutamylglycine** | OR = 0.59 [0.31 – 1.12], p = 0.107 | OR = 0.53 [0.23 – 1.27], p = 0.155 | OR = 0.81 [0.27 – 2.47], p = 0.712 |
| **Glycine** | OR = 0.65 [0.39 – 1.06], p = 0.085 | OR = 0.56 [0.28 – 1.10], p = 0.091 | OR = 0.90 [0.38 – 2.13], p = 0.819 |

N, number of individuals.Odds Ratio (OR) with their confidence interval in brackets were obtained after adjusting for age, sex, ABO blood groups and mutated gene (*ACVRL1* or *ENG*) when appropriate, under the assumption of additive genetic effects. ORs are given for 1 increase of standard deviation. Subgroup analyses by mutated gene were limited to HHT patients carrying likely pathogenic or pathogenic variants in ACVRL1 or ENG.

Using the allele effects on protein levels of the genetic instruments reported in **Supplemental Table 7**, we then evaluated whether genetically predicted plasma levels of F9 and AZI2 were associated with AVM in HHT patients. While no association was observed between genetically predicted plasma levels of F9 and AVM, higher genetically predicted AZI2 plasma levels were associated with a reduced risk of AVM (OR=0.80 [0.67–0.96], p=0.016) (**Table 3** – **Supplemental Table 18**). This association appeared to be stronger in patients carrying (likely-)pathogenic mutations in *ACVRL1* compared with those mutated for *ENG* (OR=0.70 vs 0.96), although the test for interaction between groups did not reach statistical significance (p=0.079).

We also investigated whether genetically predicted levels of the four aforementioned identified metabolites were associated with AVM. We did not observe any significant associations (**Table 3** – **Supplemental Table 18**). Nevertheless, despite being not significant, pattern of associations was consistent with MR-based metabolite results. Predicted levels of metabolites associated with decreased Sol-ENG tended to associate with increased risk of AVMs whereas predicted levels of metabolites associated with increased Sol-ENG tended to associate with decreased risk of AVMs.

Finally, the Area Under the Curve of a prediction model integrating ABO haplotypes for predicting the risk of AVMs in HHT patients was significantly higher than that of a model based only on age, sex and the mutated gene (0.764 vs 0.739, p=0.025). However, the addition of genetically predicted AZI2 plasma levels did not improve the AUC (0.770 vs 0.764, p=0.250).

## Discussion

By leveraging multi layers of omics data, including genomics, transcriptomics, proteomics and metabolomics, from publicly available summary statistics and raw individual data, this study identifies novel molecular determinants participating to the regulation of Sol-ENG plasma levels.

We identified nine independent loci showing replicated associations with Sol-ENG levels. These associations were robustly observed for Sol-ENG plasma concentrations measured using two complementary high-throughput proteomic platforms. Among these loci, the *ABO* locus was the one contributing the most to Sol-ENG plasma levels, explaining about 6% of their variability, while the remaining loci additionally explained ~4%. The lead variants at these loci were shown to be shared pQTLs for many plasma proteins, some of them being known to participate to biological pathways (eg endothelial cell biology, hematopoiesis and placenta growth) in which the role of Endoglin has been already discussed in the literature. Of note, *ABO* and *ASGR1* were the two loci exhibiting the largest pleiotropic effects as they were observed to be involved in the regulation of plasma levels of more than 300 proteins. Interestingly, the lead variant of the *ENG* locus, rs10987756, was not observed to be a pQTL for other protein beyond ENG, but was observed to be the sole Sol-ENG associated variants that module *ENG* expression in monocytes. This would indicate that this variant primarily regulates the intra cellular expression of Endoglin while all others variants would act more on secreted form of Endoglin and/or its cleavage. Consistent with this hypothesis is the observation that gene expressions at several GPI-anchor proteins (PIGG, PIGU and PIGZ) associated with Sol-ENG plasma levels. GPI-anchored proteins are enriched in specific membrane microdomains, including lipid rafts, which can influence the localization and activity of membrane-associated proteases. Metalloproteinases have been shown to mediate the cleavage of membrane-bound endoglin and promote its shedding (Vicen et al. 2020). Genetic perturbations affecting GPI-anchor biosynthesis could therefore alter membrane organization and potentially modulate the localization, activity, or interactions of endoglin sheddases.

Two proteins were identified by MR as being possibly causally associated with Sol-ENG plasma levels, FIX and AZI2. Through its role in the coagulation cascade, FIX may alter endothelial biology, a pathway we identified as enriched with Sol-ENG associated variants, and thereby regulate the cleavage of membrane-bound Endoglin into its soluble form. Also known as NF-Kappa-B-Activating Kinase-Associated Protein 1, the role of AZI2 in Sol-ENG regulation is also highly plausible. AZI2 is a protein that contributes to NF-kB activation (Fujita et al. 2003), the latter promoting *ENG* gene transcription in endothelial and immune cells, particularly under inflammatory or hypoxic conditions (Vicen et al. 2019). Conversely, Endoglin can modulate NF-κB–dependent pathways (Varejckova et al. 2017), suggesting a feedback loop between endothelial activation, inflammation, and vascular homeostasis. Of note, we observed a negative and significant correlation between *ENG* and *AZI2* in monocytes and in endothelial cells (**Supplemental Table 10**).

Our work also pinpoints to the possible causal association of plasma levels of four metabolites with Sol-ENG: Asparte to mannose ratio, Glucose-to-mannose ratio, Gamma-glytamylglycine and Glycine levels. Endoglin is a glycosylated membrane glycoprotein whose folding, stability, and shedding may depend on N-linked glycosylation pathways (Bernabeu et al. 2007). Interestingly, variations in the mannose pool (reflected by glucose-to-mannose and aspartate-to-mannose ratios) have been reported to modulate protein glycosylation efficiency in the endoplasmic reticulum (Ichikawa et al. 2014; Sharma et al. 2014), supporting the associations observed with these metabolites. In addition, γ-Glutamylglycine and glycine are intermediates of glutathione (GSH) metabolism, a central antioxidant system. As oxidative stress has been observed to regulate Sol-ENG in endothelial cells (Cudmore et al. 2007), we speculate that alterations in GSH turnover contribute to endothelial shedding of Endoglin. Of note, Glutathione Synthetase S was among the candidate expressions identified in our TWAS analysis adding another piece of support for the involvement of GSH pathway in Sol-ENG regulation.

Further works are needed to thoroughly investigate the regulatory mechanisms linking the identified molecular markers to Sol-ENG. In particular, identifying and characterizing the true functional variants underlying the detected statistical signals is essential, representing a crucial step before any accurate clinical translation can be envisaged.

By assessing the impact of these molecular findings on vascular complications in HHT patients, we observed that the ABO blood groups were significantly associated with the risk of AVMs in HHT patients. The clinical utility of ABO blood groups in the context of clinical diagnosis for HHT, especially for predicting AVM complications in HHT patients, deserves further investigations. We also observed that, independently of ABO blood groups, higher genetically determined AZI2 plasma levels were associated with a reduced risk of AVMs. It is worthy of note that this association tends to be mainly observed in patients with *ACVRL1*-related HHT in whom member-bound Endoglin is not expected to be altered, while this association was much weaker in patients with ENG pathogenic mutations and expected to be deficient in Endoglin. Measuring plasma AZI2 in this cohort would enable us to determine the full predictive value of this biomarker for AVMs, rather than just the partial and limited predictive contribution attributable to the genetic regulation of AZI2 plasma levels. In addition, the absence of plasma Endoglin and AZI2 levels data in our cohort of HHT patients prevented us from conclusively determining whether plasma AZI2 levels are associated with AVMs and whether this association is mediated by Endoglin levels. Importantly, our MR findings for AZI2 were derived exclusively from trans-pQTL genetic instruments, indicating that the observed association is likely mediated through upstream regulatory mechanisms rather than the protein itself. Consequently, direct pharmacological targeting of AZI2 may not reduce the risk of AVMs. These findings underscore the need to further characterize the biological mechanisms linking AZI2 to Sol-ENG in order to identify the causal pathways involved and to determine whether alternative therapeutic targets within these pathways may be envisaged.

All molecular determinants we found to participate to Sol-ENG regulation were identified using statistical methodologies assuming additive genetic effects. Given the high level of pleiotropy we observed for many Sol-ENG associated variants, in particular at *ABO, ASGR1* and *PLAUR* loci, it would make sense to explore more complex and synergistic interactions between loci. For example, none of the identified loci map to Matrix Metalloproteinase genes despite evidence that *MMP12* and *MMP14* are involved in the shedding of Sol-ENG (Aristorena et al. 2019; Hawinkels et al. 2010). The strongest association observed at these loci were p=0.031 and p=0.070 for *MMP12* rs505770 and *MMP14* rs11624302, respectively. This pronounced pleiotropy may also partially explain why we were unable to detect evidence for causal association between Sol-ENG levels and various diseases in which Sol-ENG have been proposed as a biomarker. GWAS on Sol-ENG according to specific genotypes at the pleiotropic loci could be a way to identify disease-causing proteins and to envisage genomics-driven drug repositioning in particular subgroups of individuals. Of note, the molecular determinants of circulating Sol-ENG investigated in our study may not necessarily reflect those regulating tissue-specific Endoglin expression and its downstream biological activity. This distinction may also partly explain why our MR analyses did not provide strong evidence supporting a causal relationship with the various outcomes investigated.

Although important for a rare disease, the size of our HHT cohort is relatively modest, limiting statistical power and the ability to detect modest genetic associations. Furthermore, our analyses focused exclusively on the presence or absence of AVMs and did not address other disease-related features, such as their number, anatomical distribution, size, severity, progression, or risk of clinical complications (e.g., bleeding). Besides, hepatic AVMs were not explored despite their potential relevance, in particular in light of the MR findings observed for FIX mainly expressed in hepatocytes. These limitations highlight the need for larger studies and further international collaborations.

In conclusion, this study provides important new insights into the regulation of Endoglin and paves the way for further research, while also highlighting novel clinical and therapeutic opportunities for improving the management of HHT. This has the potential to impact other human conditions in which Endoglin plays a role.

## Methods

### Common polymorphisms associated with Endoglin levels

To identify common polymorphisms associated with plasma Endoglin levels, we meta-analyzed GWAS summary statistics from two large-scale proteogenomics resources where Endoglin has been measured. These resources include 10,708 participants from the Fenland study (Pietzner et al. 2021) and 35,559 Icelander participants of the deCODE project (Ferkingstad et al. 2021) with Endoglin plasma levels measured using the Somalogic platform. The meta-analysis of these GWAS results was performed using the METAL software implementing the Z-score fixed-effect model (Willer et al. 2010). The heterogeneity of genetic associations across studies was assessed using the Cochran-Mantel-Haenszel method and its magnitude was expressed in terms of I^2^ (Mantel et Haenszel 1959). Only associations with moderate heterogeneity I^2^ <30% were considered.

We further conducted clumping analysis of the meta-GWAS results, based on linkage disequilibrium (LD) with a threshold of pairwise r^2^<0.01 over a distance of 10Mb. The analysis was performed using the ieugwasr R package. (10.32614/CRAN.package.ieugwasr) and the European 1000 Genomes Phase 3 reference panel. To optimize variant coverage, clumping was performed based on Hg38 positions rather than rsIDs. At the end of this analysis, SNPs which were identified as significantly (p<5×10^−8^) associated with sol-ENG plasma levels were considered as lead SNPs and kept for further downstream replication, fine-mapping and integrative analyses (see methods below).

### Transcriptome wide association study (TWAS)

From the meta-GWAS summary statistics, we conducted a transcriptome wide association study (TWAS) complemented by conditional and colocalization analyses with FUSION pipeline (Gusev et al. 2016; Giambartolomei et al. 2014), to identify and prioritize gene expressions. Inferred gene expressions from 52 tissues and cell types were analysed, and only results from LASSO models were considered. Associations were deemed significant if they reached the multiple testing-corrected threshold of P<8.3×10^−6^ based on the average number of genes tested per tissue/cell types (N=5,999). Based on colocalization and conditional analyses (Giambartolomei et al. 2014), we then prioritized results with a posterior probability of a shared causal variant (PP4) greater than 0.75.

### Mendelian Randomization

Capitalizing on the results of our meta-GWAS analysis on sol-ENG, we applied several Two-Sample Mendelian Randomization (MR) methodologies including Inverse Variance Weighted (IVW) (Burgess et al. 2013), Weighted Median (Hartwig et al. 2017), Egger (Spring et al. 1989), and MR-PRESSO (Verbanck et al. 2018) implemented in the TwoSampleMR (version 0.5.7) and MR-PRESSO (version 1.0) R packages. For these MR analyses, we used as genetic instruments for sol-ENG the independent lead SNPs identified after the clumping analysis.

#### With plasma proteins

For these analyses, we used publicly available GWAS results 2,940 proteins measured with the antibody-based Olink Explore 3072 PEA in UK Biobank study (N=34,557) (Sun et al. 2023). We only kept proteins influenced, at P<5×10^−8^, by more than two independent *cis* or *trans* genetic instruments. This led to the selection of 2,188 unique proteins present in UK Biobank. For each of these proteins, the aforementioned MR methods was applied. To account for multiple testing, we used a proteome-wide significance level of P<2.28×10^−5^ (~0.05/2188) based on the IVW method.

#### With plasma metabolites

For these analyses, we used GWAS summary statistics from 459 metabolites or metabolite ratios that were reported in (Y. Chen et al. 2023) as having at least two genetic instruments (P<5×10^−8^). As a consequence, a statistical threshold of P<1.09×10^−4^ (~0.05/459) was used to identify significant metabolite-based MR results.

#### With clinical traits

We also deployed these aforementioned MR analyses to assess a possible causal role of plasma Endoglin on preeclampsia (Tyrmi et al. 2023), systemic sclerosis (Jiang et al. 2021), coronary artery disease (Aragam et al. 2022), myocardial infarction (Hartiala et al. 2021), stroke (Mishra et al. 2022), and venous thrombosis (Thibord et al. 2022) using dedicated summary GWAS statistics.

For plasma proteins and metabolites, bi-directional MR analyses were performed. Based on these results, we selected plasma biomarkers that showed statistical evidence (using the same thresholds as those mentioned above) of a causal effect on sol-ENG levels, with no evidence supporting reverse causality.

### Replication, Fine-Mapping, and Integration Strategy

#### Replication in UK Biobank and fine-mapping in the 3C-Dijon study

We used summary statistics from the GWAS of sol-ENG levels conducted in the UK Biobank (N=34,557) (Sun et al. 2023), where plasma sol-ENG was measured using the antibody-based Olink platform, to provide additional statistical support for the genome-wide association findings obtained from the SomaLogic-based metaGWAS. By applying the FUSION pipeline to the UK Biobank summary sol-ENG GWAS, we also assessed the replication of main TWAS findings.

The lead pSNPs that were consistently associated with sol-ENG in UK Biobank were also investigated for further fine-mapping analyses using individual data from an additional plasma proteogenomic resource consisting of a sample of 1,056 population-based participants of the 3C-Dijon study (Godin et al. 2008; Debette et al. 2024) profiled on the Olink Explore 3072 panel. After exclusion of two participants presenting with extremely low outlier Endoglin values, 966 3C-Dijon participants with GWAS data remained for fine mapping analysis. In 3C-Dijon, association of common polymorphisms with sol-ENG levels was conducted on centered and standardized NPX values adjusted for age and sex. The Haplo.stats R package was also used to conduct LD and haplotype analyses of SNPs located at the same locus.

We also utilized the 3C-Dijon data to assess the biological correlation between plasma levels of sol-ENG and those of proteins identified through the protein-based MR analysis.

#### Impact of sol-ENG associated polymorphisms on ENG Monocyte Expression

To determine the impact of the identified pQTLs on *ENG* monocyte expression, we analyzed expression quantitative trait loci (eQTL) data from the Cardiogenics Transcriptomic Study (CTS), which has been previously described (Garnier et al. 2013; Heinig et al. 2010). Briefly, the Illumina Human Ref 8 version3 beadchip was used to assess RNA genome-wide expression in monocyte from 758 CTS participants in which the *ENG* gene expression was characterized by the ILMN 1760778 probe. CTS participants were also genotyped for the Illumina Sentrix Human Custom 1.2M and Human 610-Quad beadchips. Genotype data were further imputed by Mach (version 1.0.18c) software according to the 1000 Genomes Phase3V5 (Hg19) reference panel. The association of pQTL SNPs with *ENG* gene expression was tested using linear regression model, followed by haplotype analyses, conducted with the Haplo.stats R package, for SNPs located at the same locus. CTS expression data were also used to estimate the correlation between *ENG* expression and the expression of genes encoding proteins identified through the MR analysis. All statistical analysis of CTS data were adjusted for age, sex and center.

#### Transcriptomic correlates of ENG expression in endothelial cells

HUVEC were isolated from umbilical cords and pooled into six sample sets that were cultured for 0.5, 1, 2, 3, 4, 6, 8, 12, 24, 36, 48, or 72h, prior to analysis by RNAseq, as previously described (Struck et al. 2024). Pearson correlation coefficients between ENG mRNA expression levels and those of the candidate genes were calculated in R using the corr.test function from the psych package (v 1.8.4).

#### Identifying Shared Mechanisms with sol-ENG Regulation Through Plasma pQTL Analysis

For each of the lead SNPs that showed consistent effects on sol-ENG between Somalogic and Olink platforms, we retrieved their association with other plasma protein levels from the deCODE, Fenland and UK Biobank studies. We then meta-analysed their effects using the Mantel-Haenszel fixed-effects methodology (Mantel et Haenszel 1959) to identify other than proteins than ENG that are influenced by sol-ENG-associated SNPs, suggesting possible shared mechanisms. For this analysis, a significant threshold of P<8.6×10^−6^ was applied, corresponding to a Bonferroni correction for 5,812 unique proteins. Finally, we highlighted proteins influenced by several independent sol-ENG associated SNPs.

### Genetic correlation analysis

To estimate the correlation between genetically determined plasma levels of Endoglin measured by the Somalogic and Olink platforms, we used the Linkage Disequilibrium Score regression approach (B. K. Bulik-Sullivan et al. 2015; B. Bulik-Sullivan et al. 2015) implemented in the LDSC package (https://github.com/bulik/ldsc). This method was applied to Endoglin summary GWAS statistics separately from the meta-analysis of the Fenland and deCODE GWAS studies and from the UK Biobank.

### Genetic association with arteriovenous malformations in HHT patients

In this work, 760 patients with confirmed HHT associated with a pathogenic variant in *ENG* or *ACVRL1*, and followed up at the French National Reference Center in Lyon were studied. All patients with complete clinical data available in the French HHT patient database (CIROCO) and accessible DNA samples underwent genome-wide genotyping with the Illumina GSAMD.24-v3.0 array containing 730,059 SNPs. DNA samples were obtained from the laboratory responsible for the genetic diagnosis of HHT, and DNA extraction was carried out using standard procedures. The study was approved by the Scientific and Ethical Committee of Hospices Civils de Lyon (approval #23-5212). All participants gave informed written consent, in accordance with the Declaration of Helsinki.

Genotyped individuals demonstrating close relatedness, gender discordance or extreme heterozygosity were excluded (n=22) (Plink 1.90 software). SNPs showing significant (p<10^−5^) deviation from Hardy-Weinberg Equilibrium (HWE), with minor allele frequency (MAF) less than 1% or genotyping call rate <99% were filtered out. This led 738 individuals and 494,904 SNPs for the imputation step. Imputation was performed on the Helmholtz Munich Imputation server with the HRC all population panel version r1.1 2016. File preparation was done using scripts provided online (https://imputationserver-doc.readthedocs.io/en/latest/prepare-your-data/). Among remaining patients, 676 underwent screening for pulmonary and/or cerebral arteriovenous malformations and were included in the statistical association analysis. Association of lead SNPs with the presence of AVMs was tested using logistic regression model adjusted for age, sex and mutated gene (*ENG* or *ACVRL1*).

## Data Availability

Part of the data analyzed in this study is publicly available. The remaining private datasets are available from the corresponding author upon reasonable request.

https://www.omicscience.org/apps/pgwas/

https://www.ebi.ac.uk/gwas/

https://metabolomips.org/ukbbpgwas/

https://www.decode.com/summarydata/

## Acknowledgments

This project was carried out in the framework of the French National Research Agency (ANR) ANR-23-CE17-0042-01 program as part of the ENDOMORF project and of the INSERM GOLD Cross-Cutting program (D-A.T.). O.S was financially supported by a grant of the Lefoulon-Delalande Foundation. IC and FS were supported by the Digital Public Health Graduate Program (DPH), a PhD program supported by the French Investment for the Future Program (grant no. 17-EURE-0019). The Three-City (3C Study) study is conducted under a partnership agreement between the Institut National de la Sante et de la Recherche Medicale (INSERM), the Victor Segalen-Bordeaux II University, and Sanofi-Aventi. The Fondation pour la Recherche Médicale funded the preparation and initiation of the study. The Three-City study is also supported by the Caisse Nationale Maladie des Travailleurs Salariés, Direction Générale de la Santé, Mutuelle Générale de l’Éducation Nationale, Institut de la Longévité, Conseils Régionaux d’Aquitaine et Bourgogne, Fondation de France, Ministry of Research-INSERM Programme Cohortes et collections de données biologiques, the Fondation Plan Alzheimer (FCS 2009-2012), the Caisse Nationale de Solidarité pour l’Autonomie (CNSA), and Roche. The 3C proteomics project was supported by a grant overseen by the French National Research Agency (ANR) as part of the “Investment for the Future Programme” ANR-18-RHUS-0002 and by the Precision and global vascular brain health institute funded by the France 2030 investment plan as part of the IHU3 initiative under grant agreement ANR-23-IAHU-0001. The CARDIOGENICS project was funded by the European Union FP6 program (LSHM-CT-2006-037593). This work was supported by a French government grant managed by the Agence Nationale de la Recherche within the France 2030 program (ANR-21-PMRB-0007). Statistical analyses benefited from the CBiB computing centre of the University of Bordeaux.

## Author contributions

OS and DAT conceived the project. OS, CP, CER, JAS, ER, FP and BJV designed the experiments. OS, CP, CER, CD, ER, and BJV performed the experiments. GM, OS, CP, CD, ER, and BJV analyzed the data. JES, ER, FP, BJV and AG provided technical support and suggestions on the project and the experiments. SDG and MT were in charge of clinical management of HHT patients. CR, CB, MT and SDG participated in collecting and implementing the French HHT cohort. JFD, MG and RO were in charge of the production and the bioinformatic analyses of the HHT GWAS data. IC and FT performed bioinformatics analysis on the 3C study under the supervision of SD. OS and DAT drafted the paper that was further shared to co-authors who read/corrected/ and approved the final manuscript.

## Figure Legends

**Supplemental Figure 1. Haplotypes distribution derived from the rs558240 and the five polymorphisms tagging for ABO blood groups**.

Results were obtained using the LDhap tool (https://ldlink.nih.gov/?tab=ldhap).

**Supplemental Figure 2. Significant findings from the TWAS analysis on Sol-ENG followed by conditional TWAS and colocalization analyses**

