## Supplementary figures and images for "Plasma Proteogenomic Uncovers Novel Molecular Mechanisms of Soluble Endoglin and Identifies AZI2 as a new Player in Arteriovenous Malformations in Hereditary Hemorrhagic Telangiectasia"

### Supplemental Figure 1

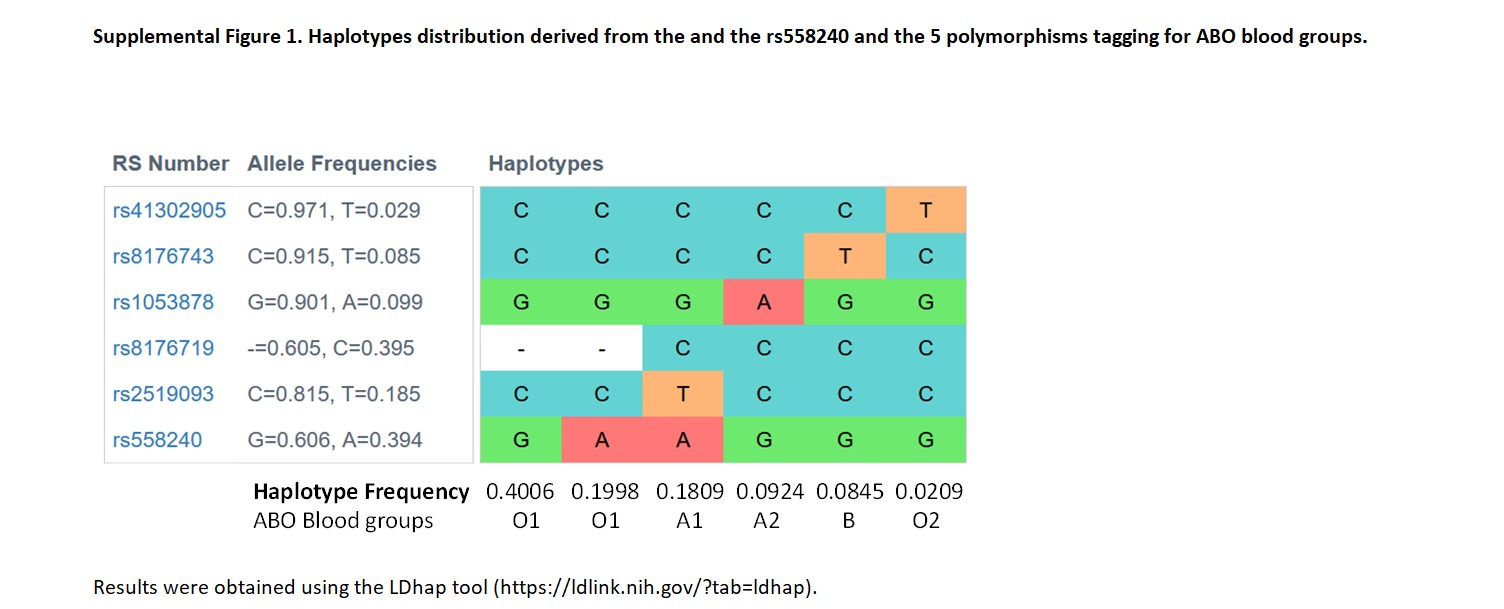

### Supplemental Figure 2

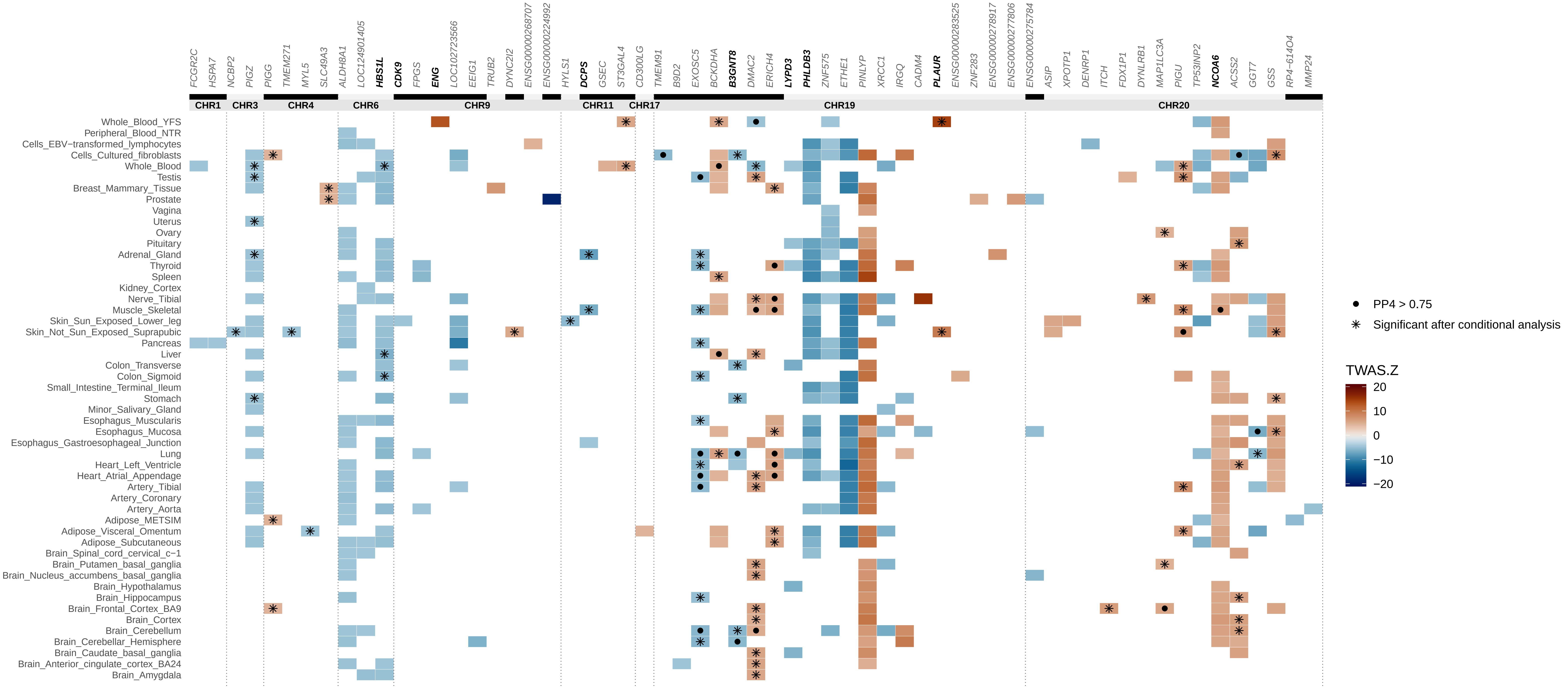


*Genes in bold were identified in the extended GWAS meta−analysis*
